# Prediction of Daily New COVID-19 Cases - Difficulties and Possible Solutions

**DOI:** 10.1101/2023.08.04.23293429

**Authors:** Xiaoping Liu

## Abstract

Epidemiological compartmental models, such as *SEIR* (Susceptible, Exposed, Infectious, and Recovered) models, have been generally used in analyzing epidemiological data and forecasting the trajectory of transmission of infectious diseases such as COVID-19. Experience shows that accurately forecasting the trajectory of COVID-19 transmission curve is a big challenge to researchers in the field of epidemiological modeling. Multiple factors (such as social distancing, vaccinations, public health interventions, and new COVID-19 variants) can affect the trajectory of COVID-19 transmission. In the past years, we used a new compartmental model, *l-i SEIR* model, to analyze the COVID-19 transmission trend in the United States. The letters *l* and *i* are two parameters in the model representing the average time length of the latent period and the average time length of infectious period. The *l-i SEIR* model takes into account of the temporal heterogeneity of infected individuals and thus improves the accuracy in forecasting the trajectory of transmission of infectious diseases. This paper describes how these multiple factors mentioned above could significantly change COVID-19 transmission trends, why accurately forecasting COVID-19 transmission trend is difficult, what the strategies we have used to improve the forecast outcome, and some of successful examples that we have obtained.

## Introduction

The World Health Organization (WHO) has declared an end to the COVID-19 global health emergency on May 5, 2023[1]. The COVID-19 pandemic has made great health damages to the peoples in the world. In the past three and half years, there were nearly 7 million people who died from COVID-19 globally[2]. Throughout the pandemic, researchers used mathematical models to analyze COVID-19 data for better understanding transmission patterns, monitoring disease severity, anticipating future epidemic outcomes[3], and justifying the adoption of intervention measures[4]. Among these mathematical models, compartmental models describing the disease as a sequence of different stages encountered upon infection to recovery, such as *SEIR* (*S*usceptible-*E*xposed-*I*nfectious-*R*ecovered) model, have been generally adopted to forecast or simulate future transmission trajectories[5, 6]. These compartmental models provide a parsimonious (i.e., using few parameters) approach to understanding important behaviors of epidemic pathways. Experience has shown that such models generate robust results that strengthen their usefulness[7]. However, it has been recognized that forecasting COVID-19 transmission trajectories is still a big challenge to the mathematical modelers[3, 5, 7, 8]. Multiple factors, such as interventions on social distancing, face masks, vaccination, emerging of new variants of COVID-19 with more contagious, may affect the accuracy of prediction from compartmental models[3, 5, 7]. Understanding how these factors affect forecast results is important to improving forecast accuracy. Furthermore, the compartmental models assume that infected individuals in each related compartment have no temporal heterogeneity, so all infected individuals in a compartment have the same probability to transfer to their next compartment. However, the realty is different: individuals are usually infected in differential days with a chronological order, so on average, individuals infected earlier in one compartment will be transferred to their next compartment at an earlier time. We recently developed a new compartmental model[9], the *l-i SEIR* Model. The letters *l* and *i* are two parameters in the model representing the average time length of the latent period and the average time length of infectious period. The *l-i SEIR* model takes into account of temporal heterogeneity or the chronological order of infected individuals. It was demonstrated that, when calculating the transfer rate of infected individuals from one compartment of the *SEIR* model to the next compartment, the temporal homogeneity approximation in the conventional *SEIR* model leads to calculation errors that increase linearly with the rate of change in the number of infectious individuals. Despite the improvement in calculation accuracy of the *SEIR* model after taking into account of the chronological order of infected individuals in the model, other factors mentioned above are still able to largely affect prediction values from the model. In this paper, we will describe the difficulties that we have encountered in predicting transmission trajectories of COVID-19 from the *l-i SEIR* model and some strategies that we have used to overcome these difficulties. Our data analysis focuses on the data of COVID-19 transmission caused by COVID-19 omicron variants in the United States.

## Methods

The *l-i SEIR* (*S*usceptible-*E*xposed-*I*nfectious-*R*ecovered) epidemic model is described by the following recursive equations:

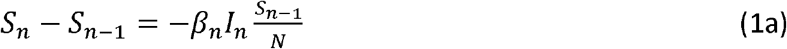

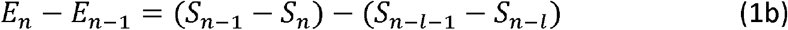

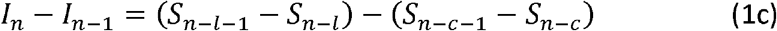

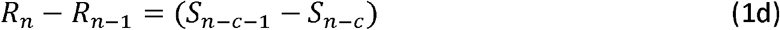

The solution of the *l-i SEIR* model in closed-form is given as below[9, 10]:

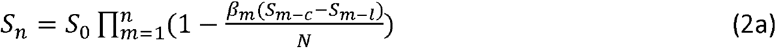

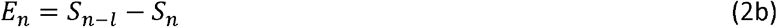

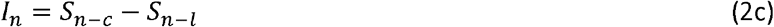

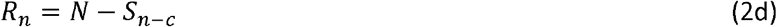

To connect the calculated model variable with the daily new COVID-19 cases, we assume:

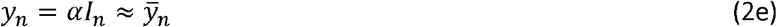

In the above equations, *S*_n_ is the number of remaining susceptible individuals who are able to contract the disease on day *n*; *E*_n_ is the number of exposed individuals who are in the latent period before becoming infectious; *I*_n_ is the number of infectious individuals who are in the infectious period and are capable of transmitting the disease; *R*_n_ is the number of recovered individuals who are becoming immune; *β*_*n*_ represents the transmission rate coefficient on day *n*; *y*_n_ represents the calculated number of the daily new cases; the coefficient *α* is a fraction between 0 and 1; *αI*_*n*_ means that only a fraction of individuals in the infectious period becomes the confirmed daily new cases; and 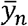 represents the reported number (7-day average) of the daily new COVID-19 cases in the United States. The parameter *c* represents the sum (*c* = *l+i*) of the average time length of latent period (*l*) and the average time length of infectious period (*i*). For the initial condition of Eqns. (2a)-(2d), we assume: (a) *S*_*n*_ *= N* and *E*_*n*_ = *I*_*n*_ = *R*_*n*_ = 0 as *n* < 0; and (b) *S*_0_*=N-*1, *E*_0_=1, *I*_0_=*R*_0_=0. Here, *N* is the number of susceptible people right before the infectious disease spreads out. If all people in the population are susceptible to the infectious agents before the infectious disease spreads out, *N* equals to the number of population *P*. However, if a portion of people has immunity to the infectious disease before the infectious disease spreads out, *N* is smaller than *P*. Eqns (2a) – (2d) were derived based on the following assumption: Change in *S*_n_ is proportional to *I*_*n*_ and proportional to *S*_*n*_/*N*. <u>This assumption implies that any person infected cannot be re-infected by the infectious agents because they have obtained immunity to the infectious agents</u>.

From Eqn. (2a), we can easily calculate all *S*_*n*_ if parameters *l* and *c*, the coefficient *β*_*n*_ and the initial conditions regarding *S*_*n*_ (*n*≤0) and *N* are known or given. From the calculated *S*_*n*_, we can further find *E*_*n*_, *I*_*n*_, *R*_*n*_ from Eqns. (2b)-(2d), and find *y*_*n*_ from Eqn. (1e) if *α* is known. We have demonstrated how to obtain the values of *l, c* and *α* from the reported daily new COVID-19 cases in the early period of the outbreak of infectious diseases[11, 12]. Thus, theoretically, we can use the *l-i SEIR* model eqns. (2a)-(2e) to predict the trajectory of COVID-19 daily new cases if *β*_*n*_ and *N* (or *S*_0_) are known and the underlying assumption (no re-infection) to the model equations remains true.

## Results and Discussion

In the past two or three years, we used the *l-i SEIR* model to simulate and predict the daily new COVID-19 cases. The first important thing being recognized was that the rate coefficient *β*_*n*_ of COVID-19 transmission varies with time. Why *β*_*n*_ of COVID-19 transmission is not a constant but varies with time?

The coefficient *β*_*n*_ can be considered as a weight factor reflecting the efficiency of interaction between *I*_*n*_ (the number of infectious people) and *S*_*n*_/*N* (the probability that the susceptible people is going to be infected) in the rate equation (1a), which determines the transmission rate of infectious diseases. Compared to the common flu, COVID-19 is highly contagious and has a relatively high mortality rate. In the early stages of the COVID-19 outbreak, when most people were not immune to the coronavirus, the death toll from COVID-19 would be very high if interventions of social distance and face mask policies were not applied to slow the spread of COVID-19 from person to person. Therefore, some interventions including social distancing and face masking were generally used to reduce COVID-19 transmission rate via decreasing the transmission rate coefficient *β*_*n*_. However, the quantitative relationship between these interventions and the value of *β*_*n*_ are unknown while the spread rate of COVID-19 in the early outbreak period is highly sensitive to *β*_*n*_. As a result, it is difficult to accurately predict at which day the peak of daily new cases will be reached and the height of the peak in the early outbreak period of COVID-19. Furthermore, COVID-19 vaccines and/or COVID-19 variants can also greatly affect the number of COVID-19 daily new cases by changing the number of remaining susceptible people *S*_*n*_ and/or the number of total susceptible people *N*, the coefficient of transmission rate (*β*_*n*_), and the ratio of reinfection as a percentage of total infections. In the following, we will describe how these problems were addressed for predicting the trajectory of COVID-19 transmission in the United States.

### Simulations and predictions of spread of COVID-19 omicron variants in the United States

After mid-December 2020, COVID-19 vaccines were given to people in the US. Since then, COVID-19 vaccines gradually became an important factor to affect the trajectory of COVID-19 transmission. In 2021, the COVID-19 alpha variant caused a transmission wave peak in mid-April and the delta variant caused a transmission wave peak in early September[13]. In this situation, multiple factors including vaccination (affect *S*_*0*_), breakthrough infection[14] (affect *S*_*n*_), reinfection[15] (affect rate equations), and intervention measures (affect transmission rate coefficient *β*_*n*_) were able to affect the trajectory of *y*_*n*_, making simulations/predictions of *y*_*n*_ trajectory more complicated because the coexistence of these factors made it difficult to identify who were susceptible and who were immune in the US. This complicated situation changed when the Omicron variant began to spread. The Omicron variant had the strongest breakthrough infectivity and re-infectivity compared to the other previous COVID-19 variants[14, 16, 17]. Vaccine effectiveness to omicron, comparing to Delta variants, dropped from 0.52 to 0.38 for those who had had their second dose 180 days earlier or more[17]. Considering that many people in the US have only received one dose or even have not received vaccines, the actual number of people with immunity to omicron variant of coronavirus may be less than 38% (0.38) of the US population (*P*=330,000,000). Our simulations show that the transmission of omicron variant in the US can be treated as the transmission of a new infectious disease from the beginning by assuming that only a fraction ∼0.25 of the US population has immunity to the Omicron original variant in the early period of Omicron-induced COVID-19 outbreak in the US. This indicates that *N* is ∼75% of the population *P* (*N*≈0.75*P* =250000000). To simulate/predict the transmission process of Omicron variants, we first estimated the value of *β*_n_ from the reported number of daily new COVID-19 cases before Omicron started to spread out in the US by using the method described previously[9, 12]. This estimated value of *β*_*n*_ was used as the initial value of *β*_*n*_ for simulating Omicron-induced daily new COVID-19 cases.

#### Prediction of peak height and peak day of reported daily COVID-19 cases 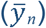 induced by Omicron variants

Usually, in the period of the COVID-19 outbreak, the number of new COVID-19 cases rapidly increases, and thus different interventions are applied for slowing down COVID-19 transmission rate, resulting that *β*_n_ gradually decreases before seeing the peak number of reported daily new COVID-19 cases 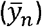. Because *β*_n_ is not a constant but decreases with time before the *y*_n_ peak passes, it is difficult to know the exact value of *β*_n_ in the future and thus it is difficult to predict the exact height and date of the *y*_n_ peak. To address this problem, we examined the magnitude of the prediction error when using the latest available number of daily COVID-19 cases 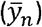 during the early rising phase of the COVID-19 outbreak to predict the height and date of the upcoming 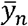 peak. In the simulations, we assumed that *l*=4, *i*=10[9, 11, 12] and *N*=250,000,000. Table 1 lists these values of *β*_n_ determined before the peak of 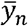 from December 25, 2021 to January 9, 2022 and the corresponding dates when these *β*_n_ values were determined. These values of *β*_n_ were determined by fitting the calculated *y*_*n*_ to the reported daily COVID-19 cases 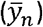 as described previously[9, 11, 12]. The *β*_n_ values determined before December 25, 2021 were not listed. The first row of the data in Table 1 shows that *β*_n_=0.4 on December 25, 2021. Supposing that we were on December 26, 2021, knowing all values of *β*_n_ up to December 25, 2021, and assuming that *β*_n_ remains constant 0.4 after December 25, 2021, we could calculate a *y*_*n*_ curve (the green line, Fig 1) from *l-i SEIR* model. From the calculated *y*_*n*_ curve, we could know the height and the date of the *y*_*n*_ peak. In this way, we forecasted the height and date of 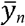 peak based on available COVID-19 data as of December 25, 2021. In the same way, we forecasted the 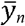 peaks and heights on other dates (Dec. 27, 29, Jan. 2, 4 and 9) listed in Table 1. The forecasted number of daily new COVID-19 cases (*y*_n_) and the reported number of daily new COVID-19 cases 7-day average 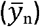 are shown in Figure 1A. The *y*_n_ peak predicted on December 25, 2021 is 1.84 million, which has the largest error comparing to the value of the reported peak 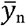 (0.81 million) on January 13 & 14, 2022. As the prediction day approaches to January 13, 2022, the predicted value of peak *y*_n_ approaches to the actual reported peak 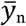 (Figure 1A). This result implies that simulations performed on earlier dates to predict the height of peak *y*_n_ may be significantly greater than the height of 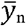 peak, whereas simulations performed on earlier dates to predict the day of the *y*_n_ peak may not produce a larger error. As shown in Figure 1B, the largest prediction error for the date of 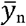 peak occurs on the day that is 15 days before the date of the reported 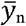 peak. This result implies that an earlier prediction day may not cause a larger prediction error for the date of 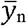 peak. Thus, by using the *β*_n_ determined from the latest reported 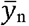 data, one can predict the date of 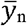 peak within a limited prediction error; and one can also predict an upper limit for the height of the 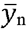 peak.

**Table 1.** The determined time-dependent *β*_n_ *N*=250000000.

| Dates | $\beta_n$ |
| --- | --- |
| Dec. 25, 2021 | 0.4 |
| Dec. 27, 2021 | 0.3 |
| Dec. 29, 2021 | 0.25 |
| Jan. 2, 2022 | 0.22 |
| Jan. 4, 2022 | 0.2 |
| Jan. 9, 2022 | 0.17 |

**Figure 1.**
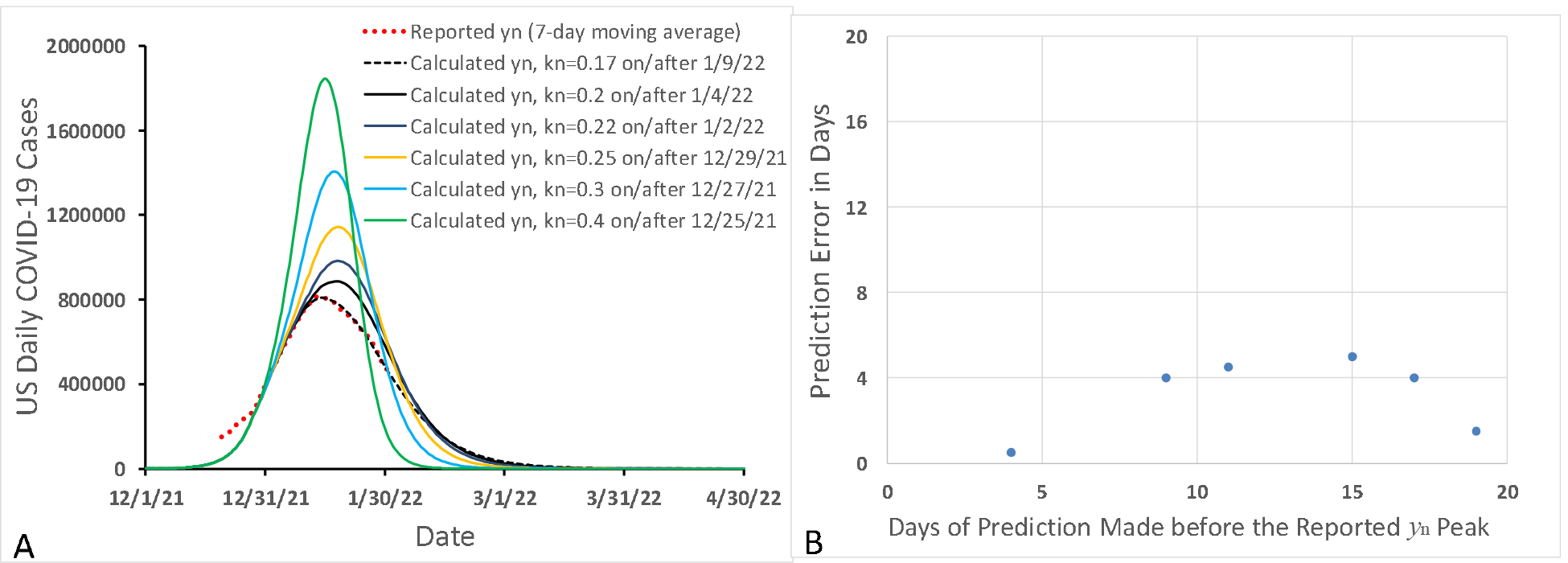
Prediction errors in forecasting peak height (A) and peak day (B) of Omicron-induced daily new COVID-19 cases in the United States.

### Prediction of lower and upper bounds of the 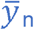curve after 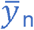peak

After the number of daily new COVID-19 cases, 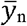, passes its peak, *β*_n_ may remain the same or even decrease a bit until it is confirmed that the peak has completely passed. Then *β*_n_ will increase because the interventions for reducing social distance and wearing face masks will be gradually lifted. Furthermore, the new sub-Omicron variants with greater infectivity may spread any day after 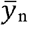 peak to increase 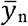 again. These unknown/undetermined factors make it almost impossible to make long-term prediction of the exact trajectory of 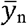. However, since *β*_n_ most likely reaches its minimum value around the 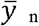 peak, if we use this minimum value of *β*_n_ to predict changes in 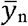 in the near future, the simulated *y*_n_ curve will be likely lower than the reported 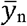 curve. This enables us to predict the lower bound of 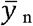 curve in the near future after the 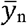 peak. The lower bound of 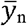 curves (dashed line) in Figure 2 was obtained by assuming that *<u>β</u>*_n_ =0.16/day after January 22, 2022. In addition to calculating the lower bound of *y*_n_ curve, we can also calculate an upper bound of the 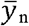 curve by assuming that *<u>β</u>*_n_ rapidly increases to 1 or a greater number in a short period of time (solid line in Figure 2). This period is chosen to be significantly shorter than the actual period needed to increase *β*_n_ in the real world. In the calculations, we assumed that no new variants or sub-variants appear in this time period to affect 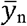 largely. As shown in Figure 2, the reported daily COVID-19 cases 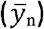 were within the predicted lower and upper bounds for 3 months, until April 22, 2023.

**Figure 2.**
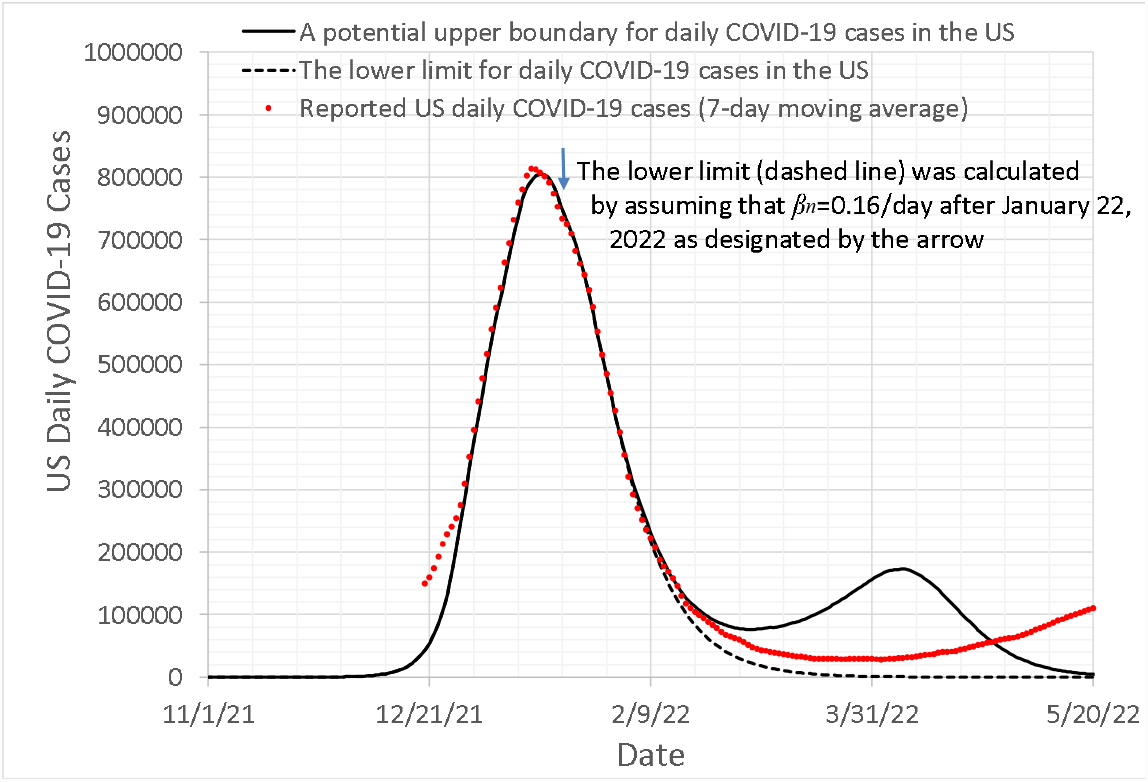
The simulated lower limit (dashed line) and upper limit (solid line) of the number of daily new COVID-19 cases (*y*_n_) induced by Omicron variants in the US after the *y*_n_ peak. The reported *y*_n_ (red dotted line) is on or between the lower and upper limits for the simulated *y*_n_.

### Simulation/prediction of Omicron sub-variants induced increases in 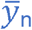

After the Omicron-induced large peak of COVID-19 transmission in the US, the number of daily new COVID-19 cases, *y*_n_, gradually decreased with time until early April 2022. Then, *y*_n_ started to increase again because of the increased *β*_n_ and transmission of multiple new sub-variants of Omicron with higher contagiousness comparing to the original Omicron variant. The major Omicron sub-variants that had significant contributions to COVID-19 transmission in the United States include (B.1.1.529 & BA.1.1), BA.2, BA.2.12.1, BA.2.75, BA.4, BA.4.6, BA.5, and (BQ.1 & BQ.1.1) [18, 19]. Among these sub-variants, B.1.1.529 and BA.1.1 were dominated in the big peak of 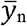 as of mid-February 2022[20], and then other sub-variants followed separately. Considering that the later Omicron sub-variants had larger infectivity, we assume that each new sub-variant mentioned above can affect COVID-19 transmission by enlarging the number of susceptible people *N*. To simulate the Omicron-induced changes in *y*_n_ after mid-February 2022, we allowed *N* to increase on some selected dates (from *N*=250,000,000 to *N*=332,400,000) between the end of 2021 and early October 2022, while *β*_n_ gradually increases to 1 as of mid-September 2022. In this way, the simulated *y*_n_ can fit the reported 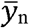 very well (Figure 3) as of Oct 23, 2022. It needs to be noted that, when *N* increases to 332400000, almost all of the population in the US has become susceptible to the highly infectious Omicron sub-variants. To predict *y*_n_ after October 23, 2022, we let *β*_n_ continuously increase to 3.5 before the end of November 2022, and remain at 3.5 after November 2022. The predicted *y*_n_ (solid line) forms a plateau from late October 2022 to the end of November 2022, and then *y*_n_ significantly decreases after early December, 2022, and *y*_*n*_ drops to nearly 1000 cases/day by the end of January 2023. This predicted result was uploaded to Twitter in late October, 2022[21]. The reported daily COVID-19 cases met the predicted results well until early December 2022[22]. Our simulation/prediction showed that after August 2022, especially after the *y*_n_ plateau in early December 2022, increasing *β*_n_ or emergence of more contagious Omicron variants would not push *y*_*n*_ up. This implies that the herd immunity to omicron has been reached in the United Sates base on the *l-i SEIR* model. In the above *l-i SEIR* model, it was assumed that any individual infected by an Omicron sub-variant would not be re-infected by any other Omicron sub-variants and any new COVID-19 variants. However, actually, Omicron-infected individuals still have a chance to be re-infected by an Omicron variant, even though the reinfection chance is very low. Therefore, the infected people are not able to form a perfect herd immunity. As we have seen, the reported daily new COVID-19 cases after late October 2022 (blue dots) formed a plateau between late October 2022 and late November 2022, which agreed with the predicted curve very well. However, the reported daily new COVID-19 cases slightly increased in the period between December 2022 and January 2023 because of the gatherings in the holiday seasons (Christmas and New Year) and a more contagious Omicron variant XBB.1.5 also appeared in this period[23]. This deviation from the predicted curve based on *l-i SEIR* model implies that a small ratio of Omicron-infected people can be re-infected by Omicron sub-variants and that the reinfection needs to be considered in the modelling.

**Figure 3.**
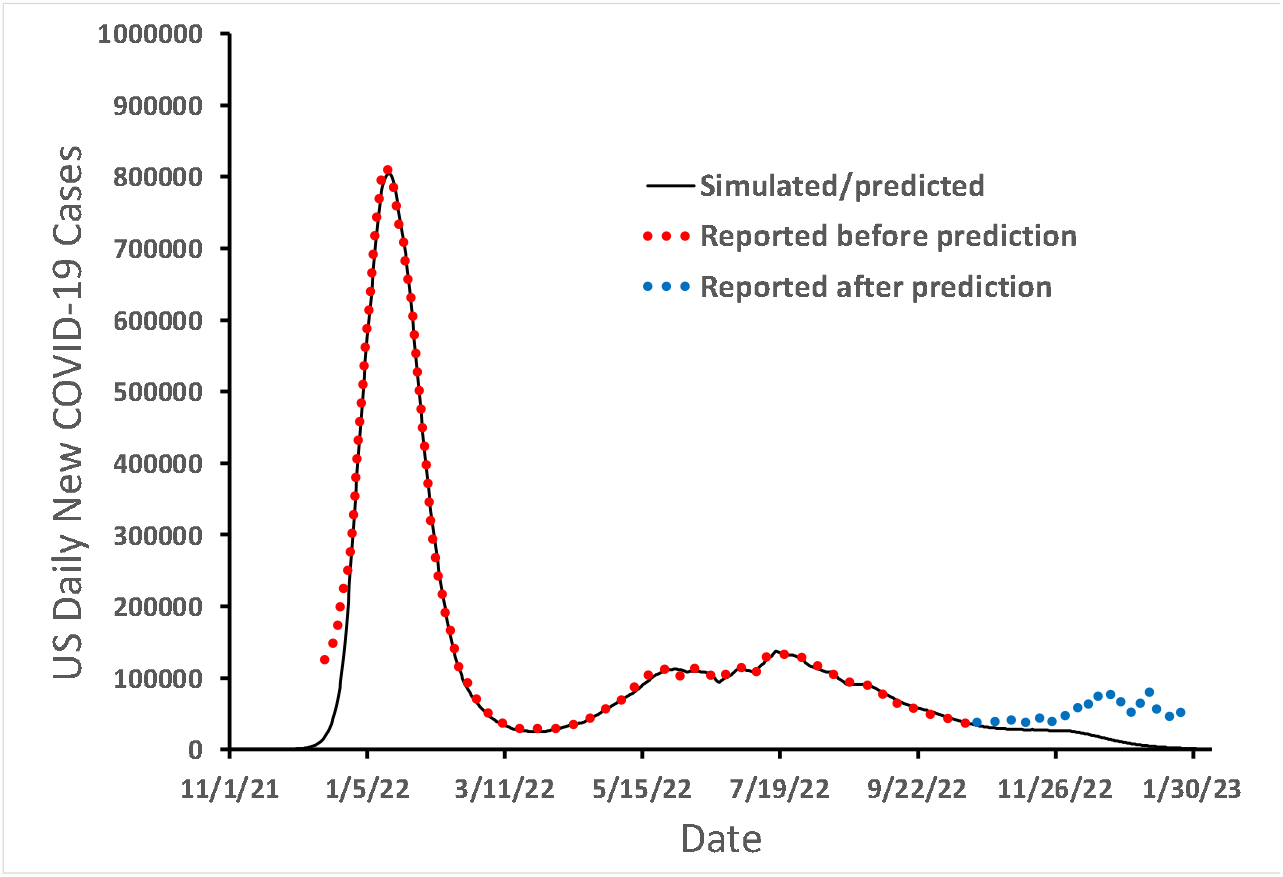
Simulated and reported number of Omicron-induced daily new COVID-19 cases in the United States without considering Omicron reinfection in the model.

### Simulation/prediction of trajectory of 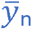 in the presence of reinfection of Omicron infections

In the above *l-i SEIR* model, the number of susceptible people *S*_*n*_ varies between 0 and *N*, or *0*≤*S*_*n*_≤*N*. If most of susceptible people have been infected, then S_*n*_ is far smaller than *N* and the ratio *S*_*n*_/*N* is near zero. Therefore, the number of daily new exposed people, (*S*_*n*_*-S*_*n-*1_) in Eqn. (1a), is also near 0. However, if the rate of reinfection of Omicron infected people is non-negligible, (*S*_*n*_*-S*_*n-*1_) must not be near zero even if all susceptible people have been infected. Thus, we suggest that in the presence of non-negligible rate of reinfection, the ratio *S*_*n-*1_*/N* in Eqn. (3a) should be replaced by [*S*_*n-*1_(≥0)*/N* + *a*_*n*_(*N*-*S*_*n-*1_(≥0))/*N*]. The second term in the square brackets consists of a reinfection coefficient *a*_*n*_ (0≤*α*_*n*_≤1) and a weight factor (*N*-*S*_*n-*1_(≥0))/*N*. When *S*_*n*_ is close to *N*, the weight factor is close to 0 and the first term in the square brackets plays the main role. However, when *S*_*n*_ is close to 0, the weight factor is close to 1 and the second term in the square brackets plays the main role. If *a*_*n*_=0, it means no reinfection. In this situation, *S*_*n*_ can vary from *N* (no one is infected) to 0 (all susceptible people are infected). If reinfection rate is non-negligible, then *a*_*n*_ is greater than 0. In this situation, *S*_*n*_ can vary from *N* (no one is infected) to a negative number. The negative number means that not only all susceptible people are infected, but also a portion of them are re-infected. Thus, in the presence of reinfection, Eqn. (1a) is replaced by the following equation:

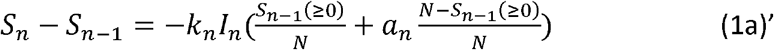

In Eqn. (1a)’, *S*_*n*_ (≥ 0) = *S*_*n*_ if *S*_*n*_>0, and *S*_*n*_ (≥ 0) = 0 if *S*_*n*_≤0. Based on Eqns. (1a)’ and (1b)-(1d), we simulated/predicted daily new COVID-19 cases on February 10, 2023[24], and compared them with reported data until May 5, 2023 (Figure 4)[25] when data of daily COVID-19 cases in the US were not updated anymore on websites. The red dots in Fig 4 represent the number of daily COVID-19 cases 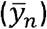 reported before the simulated/predicted trajectory of *y*_*n*_ (solid black line) was generated. The blue dots in Fig 4 represent 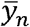 reported after the simulated/predicted trajectory of *y*_*n*_ (solid black line) was generated. The result in Fig 4 shows that the reported 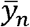 matches very well with the predicted trajectory of *y*_*n*_.

**Figure 4.**
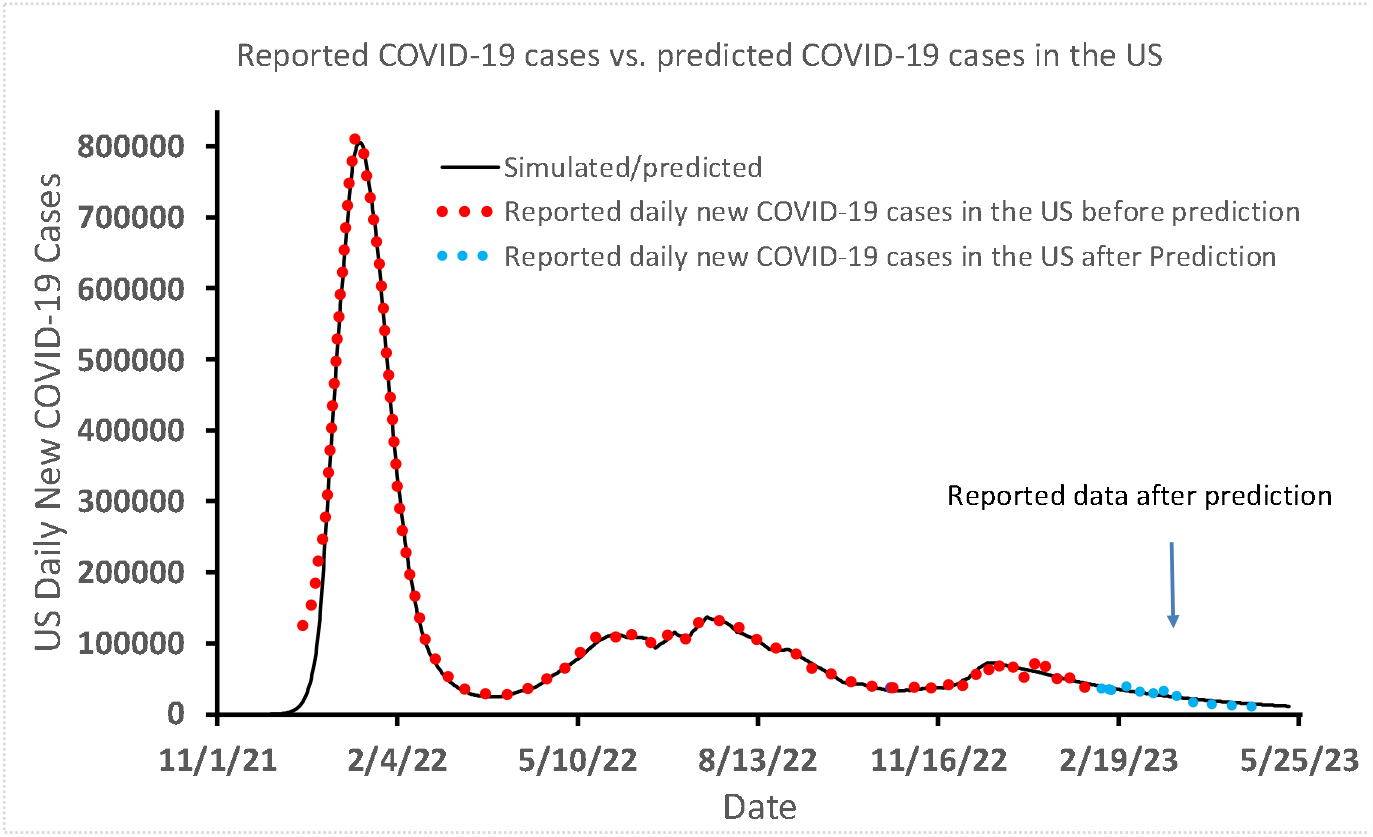
Simulated/predicted and reported number of Omicron-induced daily new COVID-19 cases in the United States after considering Omicron reinfection in the model.

## Summary

Based on *l-i SEIR* model, the author described difficulties and discussed possible solutions in forecasting the peak day and the peak height of daily new COVID-19 cases 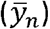, the trajectory of 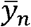 after the 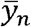 peak, and the trajectory of 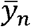 after the herd immunity was reached in the presence/absence of reinfection. Our simulations show that by using the *β*_n_ determined from the latest reported 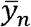 data, one may predict the date of 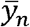 peak within a limited prediction error, and also predict an upper limit for the height of the 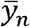 peak. It is possible to accurately predict the trajectory of *y*_*n*_ after the 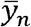 peak for a few of weeks (up to 4 weeks from 1/22/2022-2/19/2022 as shown in Figure 2) with a constant *β*_n_. However, by calculating a lower limit and an upper limit of the *y*_*n*_ curve, one may successfully predict the trace of 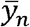 within the range between the lower limit and upper limit of the *y*_*n*_ curve for more than 3 months (from 1/22/2022 to 4/28/2022 in Figure 2). The *l-i SEIR* model without considering Omicron reinfection could not explain the remaining non-negligible number of daily new COVID-19 cases after the herd immunity was reached (*S*_*n*_/*N*≈0), suggesting that the Omicron reinfection should be taken into account in the model. The simulated *y*_*n*_ curve based on the *l-i SEIR* model considering Omicron reinfection can fit very well with the numbers of reported COVID-19 cases after the herd immunity has been reached, and the predicted *y*_*n*_ curve is in good agreement with the number of daily new COVID-19 cases reported to date May 10, 2023, twelve weeks after the prediction of *y*_*n*_ curve was made on February 10, 2023.

## Calculation Methods and Programs

### Writing codes in Excel for calculating *S*_*n*_, *E*_*n*_, *I*_*n*_, *R*_*n*_ and *y*_*n*_

We have described in detail how to put *l-i SEIR* model equations into Excel[26] to calculate model variables *S*_*n*_, *E*_*n*_, *I*_*n*_, *R*_*n*_, and *y*_*n*_ assuming that *β*_*n*_=1 and the parameters or coefficients *l, i* (or c) and *α* are known. Below is the procedure for using Eqns. (2a)-(2e) to calculate these model variables in Excel.

1. In cells B1, C1, D1, E1, F1 (row 1 of the Excel sheet shown in Table 2), write *S*_0_, *N, l, c* and *α*, respectively.
2. In cells B2, C2, D2, E2, F2 (row 2), input 329999999, 330000000, 4, 14 and 0.0145349, respectively. With these arrangements, we can easily know that *S*_0_=329999999, *N*=330000000, *l*=4, *c*=14 and *α*=0.0145349. The values of these parameters were used for analyzing COVID-19 data reported from the United States[11, 12].
3. Write variable names in row 3. Especially, column A is used for date (A3: Date); column B for the number of susceptible individuals (B3: *S*_n_ Susceptible individuals), column C for the exposed individuals (C3: *E*_n_ Exposed individuals), column D for the infectious individuals (D3: *I*_n_ Infectious individuals), column E for the recovered individuals (E3: *R*_n_ Recovered individuals), column F for the calculated US daily new COVID-19 cases (F3: *y*_n_ the calculated US daily new cases), and column G for the transmission rate coefficient (G3: *β*_n_ in US). In this Excel calculation program, we assumed that the first infection appeared on 2/2/2020[12], which is in row 28. We let *n*=0 on 2/2/2020.
4. Input dates in Excel in chronological order. First, we input 1/9/2020 in cell A4, and then click, hold, and drag the fill handle (a small black box on the bottom right) down until all of the cells we want to fill are selected. Release the mouse to fill the selected cells with the dates in chronological order as shown in column A of the Excel sheet (Table 2).
5. Set initial conditions for n≤0. Input 330000000 (*S*_n_=*N* for *n*<0) or type =$C$2 into cells B4-B27 (corresponding to dates from 1/9/2020 to 2/1/2020 (A4-A27) because our simulations suggested that the first infection in the US appeared on 2/2/2020)[12]. This operation is easy to perform if the fill handle is used. Here $C$2 is the absolute cell reference to cell C2. When the absolute cell reference is copied from one cell to another cell, the absolute cell reference will remain unchanged. Because the value of the number in C2 is 330000000, we will see 330000000 in all cells B4-B27 if B4-B27 contain =$C$2. In this case, if we change the value in cell C2, the value in all cells B4-B27 will change accordingly. Input 0 to cells C4-C27 (*E*_n_ for n<0), cells D4-D27 (*I*_n_ for n<0), E4-E27 (*R*_n_ for n<0), and F4-F27 (calculated US daily new cases *αI*_n_ for n<0). Input 1 (*β*_*n*_) to column G starting from G4 to all other cells in column G, corresponding to all dates available in column A. In row 28, n=0 and the date is 2/2/2020. The first exposed individual appeared on this day[12], so the number of susceptible individuals at n=0, *S*_0_, is 329999999. Thus, we input 329999999 or =$B$2 in cell B28. The number of exposed individuals at n=0, *E*_0_, is 1, so we input 1 in cell C28. However, *I*_0_, *R*_0_ and *αI*_0_ are still 0 at n=0, so we input 0 in cell D28, E28 and F28. So far, we have set or input all parameters and initial values required for calculating *S*_n_, *E*_n_, *I*_n_, *R*_n_ and *αI*_n_. In the following, we will input Eqns. (2a)-(2e) into row 29 in the Excel sheet to calculate *S*_n_, *E*_n_, *I*_n_, *R*_n_ and *αI*_n_ for n>0.
6. From Eqn. (2a) we know that the equation for *S*_1_ can be simplified as below:

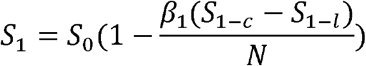

Similarly, *S*_2_ and *S*_n_ can be written as:

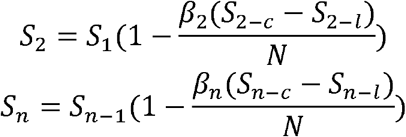

In our Excel program sheet, we let n=0 in row 28. Thus, row 29 will be n=1 and row 30 will be n=2 and so on. The value of *S*_0_ is placed in cell B28, so *S*_1_ will be placed in cell B29, and *β*_1_ placed in G29. Furthermore, we have known that *c*=14, and *l*=4[11, 12], therefore *S*_1-*c*_ or *S*_-13_ will be placed in cell B15, and *S*_1-*l*_ or *S*_-3_ will be placed in B25. Thus, to input the above equation for *S*_1_ in cell B29, we need to type the following codes (Excel codes) and click the Enter key:

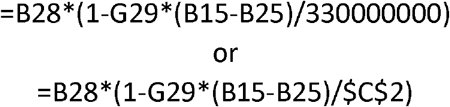
7. From Eqn. (2b) we know that the equation for *E*_1_ can be simplified as below:

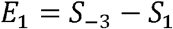

We have known in step 6 that *S*_-3_ is placed in cell B25 and *S*_1_ is placed in cell B29, so we should type the following codes in cell C29:

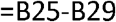
8. From Eqn. (2c) we know that the equation for *I*_1_ can be simplified as below:

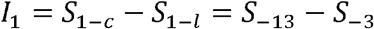

The value of *I*_0_ is placed in cell D28, so *I*_1_ is placed in cell D29. We have known in step 6 that *S*_-13_ is placed in cell B15, and *S*_-3_ is placed in B25, so we should type the following codes in D29:

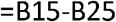
9. From Eqn. (2d) we can get the following equation for calculating the number of recovered individuals at *n*=1:

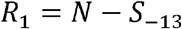

Because *S*_-13_ is placed in cell B15, so we should type the following codes in E29:

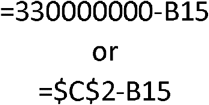
10. From Eqn. (2e) we can get the following equation for calculating the US daily new COVID-19 cases at n=1:

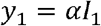

The calculated US daily new COVID-19 cases *y*_1_ is placed in cell F29. Thus, we need to type either line of the following codes in cell F29:

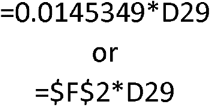

The above Excel codes mean that the value of *I*_1_ is in cell D29 and the value of *α* is in cell F2. We can directly input the value of *α*, 0.0145349, into F29 or input the absolute cell reference of *α*, $F$2, into F29.
11. So far, we have finished the Excel program for calculating *S*_1_, *E*_1_, *I*_1_, *R*_1_ and *y*_1_. To calculate any other *S*_n_, *E*_n_, *I*_n_, *R*_n_ and *y*_n_ for n>1, we just need to copy cells B29, C29, D29, E29 and F29, then paste them to B30, C30, D30, E30 and F30, and any other rows below row 30. This can be easily done by selecting cells B29, C29, D29, E29 and F29, and then drag the fill handle down to any row we wanted. If we successfully finish these steps, we should see the following Excel codes in B30, C30, D30, E30 and F30. B30 (*S*_2_): =B29*(1-G30*(B16-B26)/330000000) or =B29*(1-G30*(B16-B26)/$C$2) C30 (*E*_2_): =B26-B30 D30 (*I*_2_): =B16-B26 E30 (*R*_2_): =330000000.-B16 or =$C$2-B16 F30 (*y*_2_): =0.0145349*D30 or =$F$2*D30

**Table 2.**
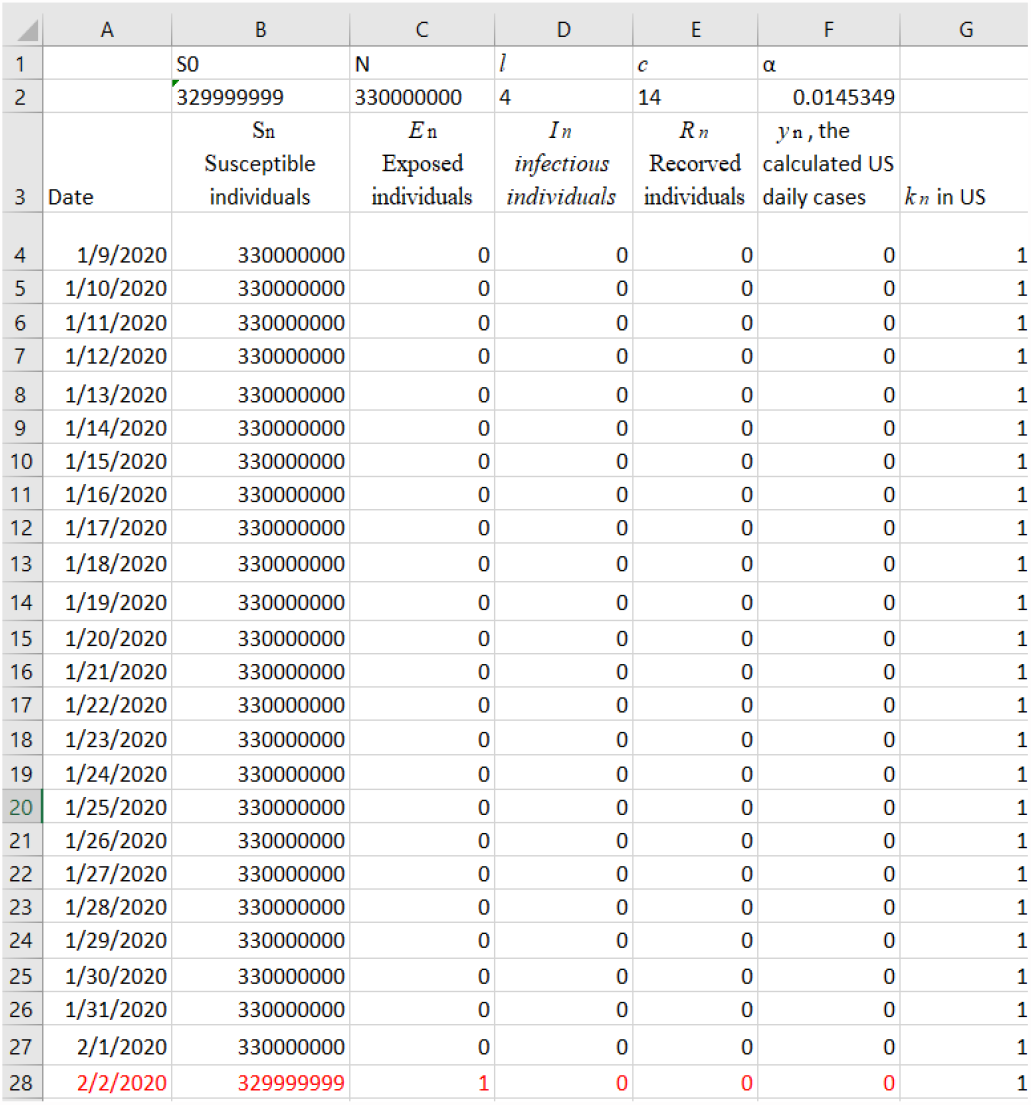
Writing parameters, naming variables and setting initial conditions in Excel.

It can be seen that all row labels increase by 1 after copying cells in row 29 to cell in row 30, but the constant 330000000, the absolute reference $C$2 and $F$2 remain unchanged. Therefore, by simply copying & pasting operation in Excel, we are able to calculate all *S*_n_, *E*_n_, *I*_n_, *R*_n_, and *y*_n_ from Eqns. (2a)-(2e), assuming that *β*_n_ is a constant (*β*_n_ =1 in this example). However, in real situation, *β*_n_ varies with time because social interventions are applied for slowing down spread rate of COVID-19. By regulating *β*_n_ for fitting the calculated *y*_n_ to the reported 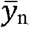, we can determining *β*_n_ for each n.

## Data availability statement

All data used in this study are publicly available.

